# HIV VIRAL SUPPRESSION AND ASSOCIATED FACTORS AMONG CHILDREN AND ADOLESCENTS ON A DOLUTEGRAVIR (DTG) BASED ANTIRETROVIRAL REGIMEN IN TANZANIA MAINLAND

**DOI:** 10.1101/2023.05.17.23290106

**Authors:** Abdallah Abtwalibe Maghembe, Marion Sumari-de Boer, Godbless Marikias, Caroline Amour, Michael Johnson Mahande

**Author notes:** Corresponding author: Abdallah Abtwalibe Maghembe, KCMUCo, P. O. Box 2240, Moshi, Tanzania.

## Abstract

**Introduction:** Antiretroviral therapy (ART) reduces morbidity and mortality due to human immunodeficiency virus (HIV) infection. The complexity and time-consuming processes, particularly in drug approvals, have contributed to a major challenge to the ongoing success of antiretroviral treatment programs among children and adolescents. In 2019, Tanzania adopted DTG as a first-, second-line and third-line treatment for CALHIV on ART after being approved by the World Health Organisation (WHO). DTG treatment has highly potent antiviral activity, a high genetic barrier to resistance, and a high safety profile. This study aimed to determine HIV viral suppression and associated factors among CALHIV on DTG-based ART in Tanzania Mainland.

**Methods:** This was a retrospective cohort analysis among children and adolescents living with HIV who were on a DTG-based regimen in Tanzania Mainland between 2019 and 2021. The study utilized routinely collected data from Tanzania Care and Treatment Centres (CTC). We analysed data using STATA version 15 software. We calculated the prevalence of viral suppression by taking the number of children and adolescents with <1000 copies/ml overall study participants. A mixed effect generalized linear model with Poisson distribution and log link function with robust estimator determined the factors associated with HIV viral suppression on a DTG-based regimen.

**Results:** A total of 63,453 CALHIV on a DTG-based regimen were analysed. The proportion of viral suppression was 91.64%. Overall, 66.19% of previously unsuppressed individuals became suppressed and 88.45% of previously suppressed remained suppressed. Factors leading to lower chances of viral suppression were age 10-14 years (aRR: 0.98; 95%CI: 0.97-0.99), previously unsuppressed prior to starting DTG (aRR: 0.92; 95%CI: 0.91-0.93), duration on ART more than 24 months (aRR: 0.96; 95%CI: 0.94-0.97), not retained in care (aRR: 0.83; 95% CI: 0.77-0.89), severe malnutrition (aRR:0.77; 95%CI: 0.69-0.94) and coastal zone (aRR: 0.98; 95% CI: 0.96-0.99), while those in WHO stage I (aRR: 1.03; 95%CI: 1.01-1.04) and ever received a multi-month prescription (aRR: 1.25; 95% CI: 1.23-1.28) had a higher chance of viral suppression.

**Conclusions:** The findings support the broad use of DTG-based regimens for eligible CALHIV. Especially those in baseline WHO stage I and those who received the multi-month prescriptions were more likely to achieve viral load suppression. Programs should improve strategies to maintain CALHIV retention in care with interventions like the promotion of teen clubs and teams.

## Introduction

HIV/AIDS is one of the greatest public health challenges in the world, whereby sub-Saharan Africa remains the region most heavily affected in the world. An estimated 1.8 (1.2-2.2) million children aged 0-14 were living with HIV at the end of 2020, and 150,000 children were newly infected. An estimated 100,000 children died of AIDS-related illnesses [1].

In Tanzania, it is estimated that around 110,000 (93,000 – 130,000) children aged 0-14 years and 1.6 (1.5 - 1.8) million adults were living with HIV/AIDS in 2020 [2]. From the Tanzania 2020 estimates, there were 6,500 new infections among children below 15 years of aged and 78% of children living with HIV are on ART [2]. The prevalence of HIV according to the THIS of 2016/2017 was less than 2% among 15-19 years for both males and females and increases with age for both sexes. HIV care and treatment programs have scaled up rapidly in many countries in sub-Saharan Africa in recent years. The National AIDS Control Programme (NACP) in Tanzania has been coordinating the scale-up of quality HIV care and treatment services at all health facility levels: including public and private hospitals and in health centres and dispensaries throughout the country, whereby 6,206 (2103 CTCs & 4103 Option B+) health facilities started to provide care and treatment by December 2018 [3].

In Tanzania, according to the Fourth Health Sector HIV and AIDS Strategic Plan (HSHSP IV) strategic outcomes, all children living with HIV under 15 years should be initiated ART and 90% of children should be retained on ART by 2022 as well as 95% of adolescents living with HIV should be retained on ART by 2022 [4]. Viral suppression is a good long-term outcome for survival and maturing to the ongoing success of antiretroviral treatment programs in limited resources countries among children living with HIV [5].

To meet the United Nations AIDS (UNAIDS) goals the agreement is to eliminate HIV by 2030 with the recommended 95-95-95 targets (diagnosis of 95% of HIV-infected people, provision of treatment for 95% of people diagnosed with HIV, and viral un-detectability in 95% of treated people). The complexity of the treatment regime among children and adolescents is still a big challenge for HIV programs in sub-Saharan Africa and forms an obstacle to the attainment of the second and third targets of the UNAIDS 95-95-95 HIV treatment targets among children and adolescents as it affects sustainable treatment and virological success of children and adolescents living with HIV accessing care and treatment service [6].

Major improvements in access to treatment have been greater for adults than for children due to the fundamental challenge in the development and approval of paediatric dosing and formulations for age and weight-specific thresholds for children and adolescents lagging behind for years and children did not benefit compared to adults despite the same level of engagement in terms of ART uptake [7].

Dolutegravir (DTG) is an Integrase Strand Transfer Inhibitor (INSTI) used as a first-line, second-line, and third-line HIV treatment regimen. According to the World health organization (WHO) updated antiretroviral therapy guidelines of 2018, Dolutegravir (DTG) is recommended at ≥6 years of age or weight ≥20 kg, and children weight for age below <20 kg use Lopinavir/ritonavir with or without being exposed to the Nevirapine among exposed infants from the risk mothers. Tanzania adopted DTG in 2019 among children and adolescents weighing ≥20kg. Hence, there is a need to study treatment outcomes of DTG-based ART regimens due to its potent antiviral activity in reducing the HIV incidence by promoting rapid viral suppression, a high genetic barrier to resistance and better safety profile [8]. Yet, in our setting, the prediction of viral suppression and associated factors has not been validated. Thus, there is still little support for the use of DTG-based regimens in mass treatment programs. Hence, there is need to assess viral suppression among children and adolescents that are on these new antiretroviral drugs. This is because a single study on DTG has shown adolescents born in Sub-Saharan countries were more likely to have virological failure compared to those born in other countries [9].

## Methods

### Study design and setting

This was a retrospective cohort study among children and adolescents living with HIV who were on an ART-initiated DTG based regimen in Tanzania Mainland between January 2019 and December 2021. The study utilized routinely collected data of Tanzania Care and Treatment Centres (CTC).

### Study population and Period

The analysis includes children and adolescents living with HIV (CALHIV) on a dolutegravir-based regimen, who were aged 1 year to 19 years in care and treatment clinics (CTCs) in Tanzania Mainland between January 2019 and December 2021. We excluded all HIV-positive children and adolescents with less than six (6) months on ART accessing care and treatment, and missing viral load result after initiation of DTG from the study.

### Sample size estimation

A total of 230,310 Children and Adolescents Living with HIV (CALHIV) patients in Tanzania mainland enrolled from January 2019 and December 2021. A total of 96,648 (41.96%) of CALHIV were on a dolutegravir (DTG) based regimen and 133,662 (58.04%) were on NNRTI and PI. We excluded a total of 33,195 (33.01%) individuals on DTG from the analysis as they did not meet eligibility criteria. The final sample was 63,453 as shown in our flow diagram (Figure 1).

**Figure 1:**
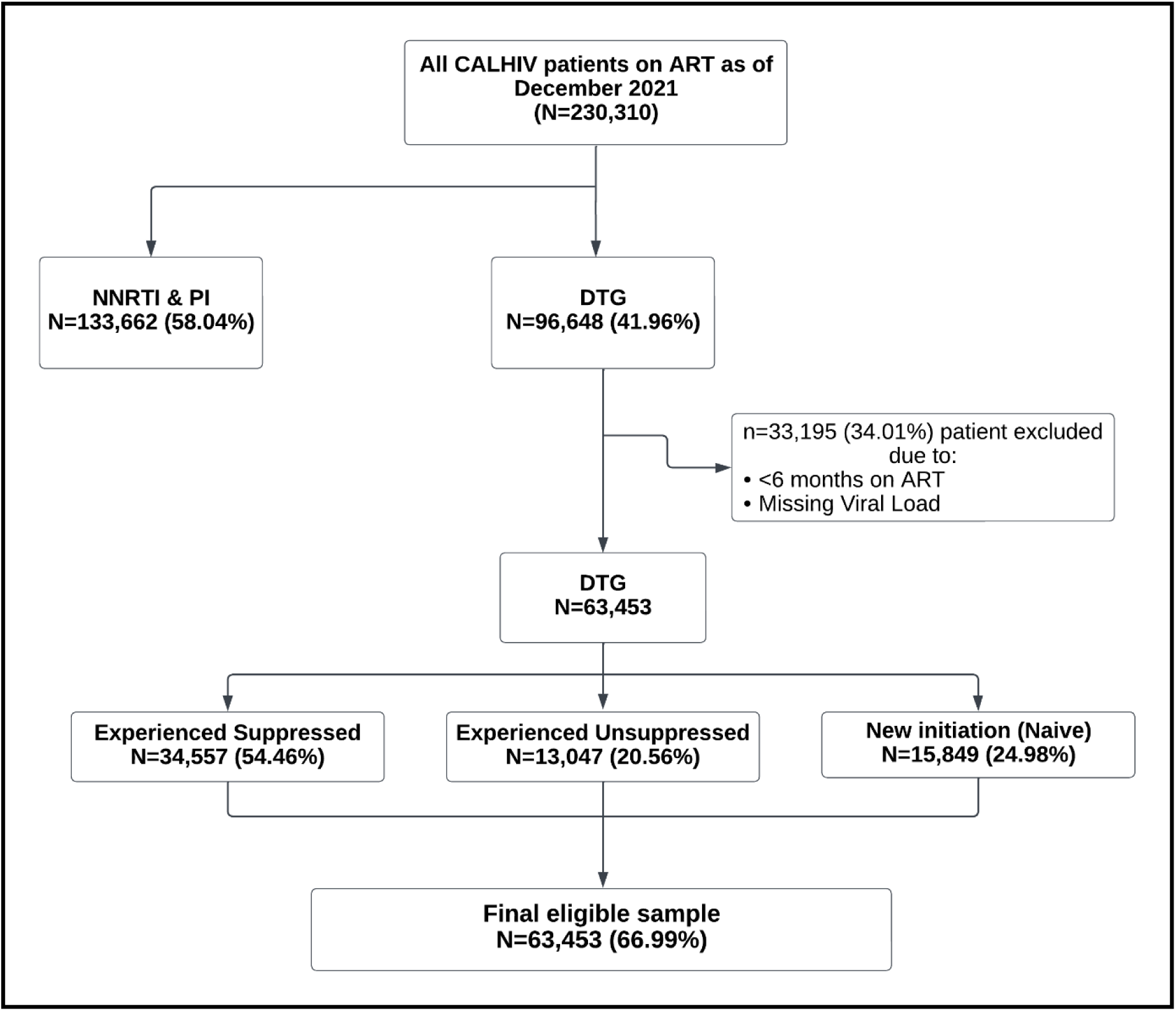
Flow chart showing the recruitment of the study population. Ethical clearance. Ethical approval with clearance number PG 12/2021 to conduct the study was obtained from KCMU College and Research Ethics Review Committee. The study utilized secondary data from routinely collected data of the care and treatment clinics (CTC) for HIV-positive clients in Tanzania hence the permission to carry out the study was obtained from National AIDS Control Programme (NACP) to access patient level information in the CTC 2 Databases. The obtained information was kept confidential and participant codes from CTC identity numbers were used instead of names.

### Data sources and study variables

This study utilized data of HIV-positive children and adolescent’s clients that were on DTG from January 2019 to December 2021. We obtained data from routinely collected data of the care and treatment clinics (CTC) for HIV-positive clients in Tanzania. We only included health facilities who initiated DTG-based regimens. The data set originates from the National Aids Control Programme (NACP).

We defined the dependent variable, HIV viral suppression, as having a last plasma viral load of less than <1000 copies/ml, which we named “Yes” 1 and having a plasma viral load of equal and greater than 1000 copies/ml we named “No” 0.

Independent variables were age in the following categories: sex, WHO clinical stages (Stage I, II, III and IV), CD4 cell count (<200, 200-350 and >350), BMI categorized as underweight (<18.5kg/m^2^), normal weight (18.5-24.9 kg/m^2^), overweight (25-29.9 kg/m^2^) and obese (≥30 kg/m^2^), prior DTG status (Experienced suppressed, experienced unsuppressed and Naive), duration on ART (6-12 months, 13-24 months and >24 months), ever received Multi Month Subscription (Yes and No), history of TB treatment (Yes and No), retention in care (In care, lost to follow up, transferred and died) and geographical zones (central, lake, western, northern, coastal and southern highland).

### Data Analysis

We analysed data using STATA version 15.0 software (Stata Corp, 2017. Stata Statistical Software: Release 15 College Station, USA). We conducted descriptive statistics for all variables; we summarized continuous variables using mean (SD) or median (IQR), depending on their distribution, and frequency and proportion for categorical variables. We computed a proportion of HIV viral suppression as the number of children and adolescents with <1000 copies/ml over all study participants. We did Chi-square tests for each explanatory variable to compare the proportion of HIV viral suppression on DTG based regimen. We used a multilevel mixed effects Generalized Linear Model with a Poisson distribution and log link function with a robust estimator to determine the association between exposures and HIV viral suppression on DTG based regimen. We interpreted the magnitude of association using an adjusted relative risk and 95% CI, while a p-value of less than 0.05 was statistically significant. We included those variables identified based on prior and clinical knowledge of the study setting in the adjusted model.

## Results

### Proportion of CALHIV on DTG-based ART

A total of 96,648 CALHIV were on a DTG based regimen from January 2019 to December 2021, representing 41.96% of all CALHIV on ART regimens. Among the DTG cohort, 17,925 (18.56%) were new ART initiations started on DTG (Naive), 52,275 (54.07%) were switched from an NNRTI and 26,448 (27.37%) were switched from a PI regimen.

### Socio-demographic and clinical characteristics among the CALHIV

We studied a total of 63,453 children and adolescents living with HIV on DTG based regimen. Findings showed that the majority of CALHIV, 21,852 (34.44%), were aged 1-4 years and the mean age was 7.74 (SD: 5.37) years at baseline. Most study participants (36,108 (56.91%)) were females. At baseline, many of the study participants were experienced suppressed with 34,557 (63.37%) prior to starting DTG, 13,041 (23.91%) were experienced unsuppressed prior to starting DTG and 6,933 (12.71%) were naïve prior to starting DTG. Most of the study participants 53,512 (84.33%) had more than 24 months duration on antiretroviral therapy. With reference to study participants, 26,977 (42.73%) were attending hospital facility type, 48,070 (76.14%) were attending a public facility and 17,152 (27.17%) were living in the lake zone. More results are shown in **Table 1**.

**Table 1:**
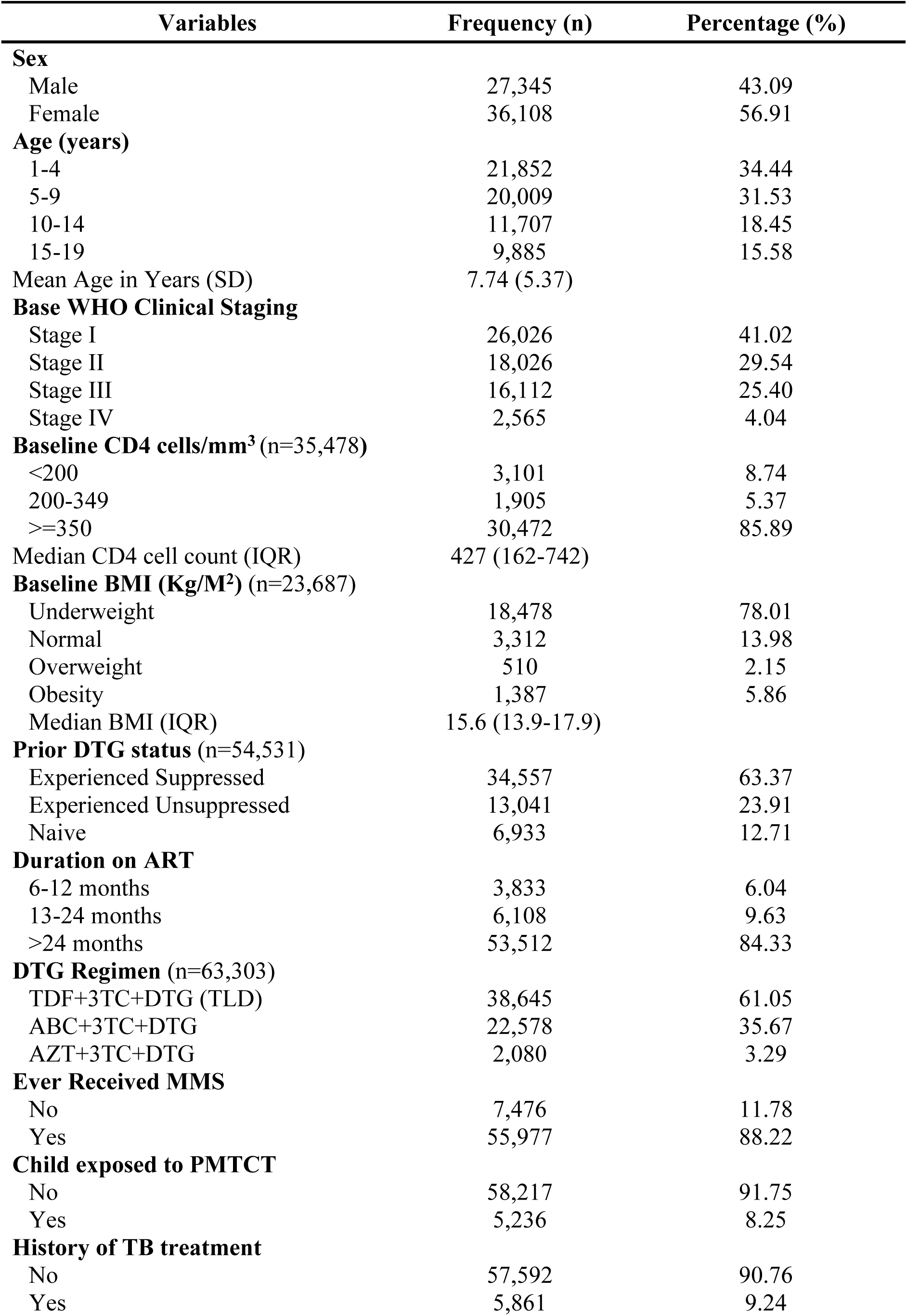
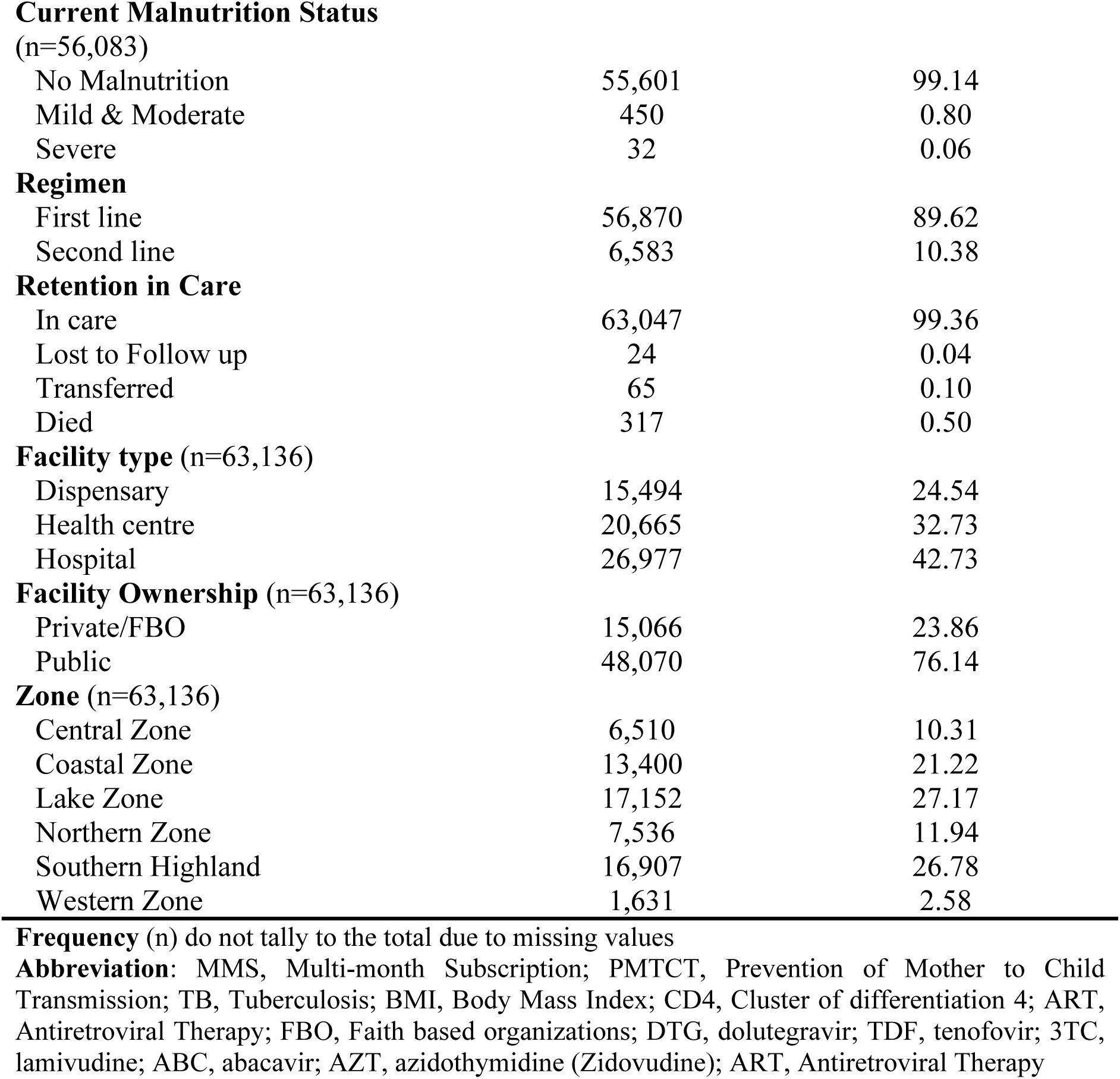
Socio-demographic and clinical characteristics in Children and Adolescent.

### Proportion of HIV viral suppression among children and adolescents on Dolutegravir (DTG) based ART

Among CALHIV on a DTG-based regimen with pre- and post-viral load tests, 88.4% of those who were suppressed before DTG, remained suppressed. Of those who were unsuppressed before DTG, 66.1% became suppressed. After six months on DTG, among naïve patients on DTG based regimen, 91.7% achieved viral suppression as shown in **Figure 2**.

**Figure 2:**
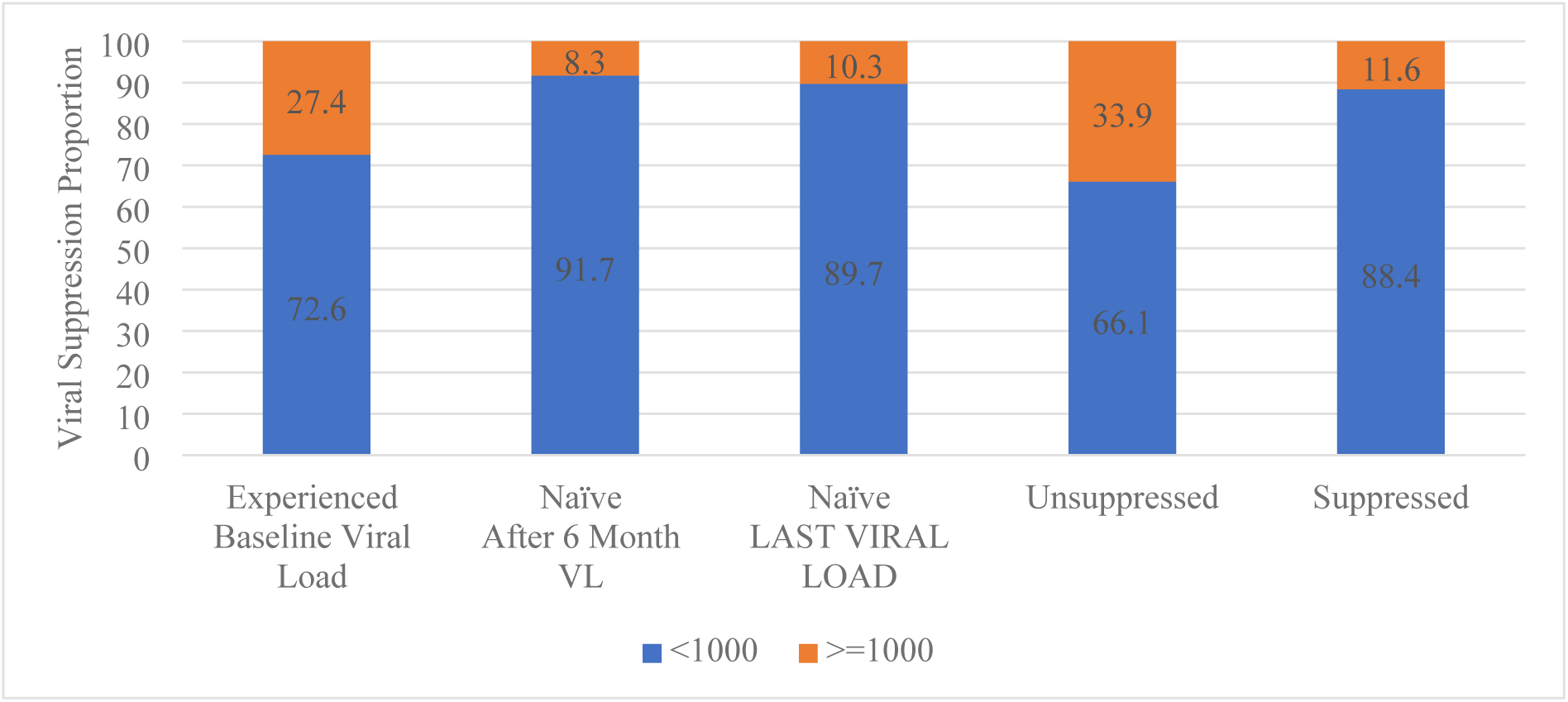
Viral Suppression at baseline, after 6 months among Naïve and last viral load among CALHIV on DTG-based regimen (N=54,531)

In addition, the proportion of CALHIV on a DTG-based regimen who achieved viral suppression was higher in ART naïve (93.2%) than in treatment-experienced CALHIV with (91.4%) p-value <0.001, as shown in **Figure 3**.

**Figure 3:**
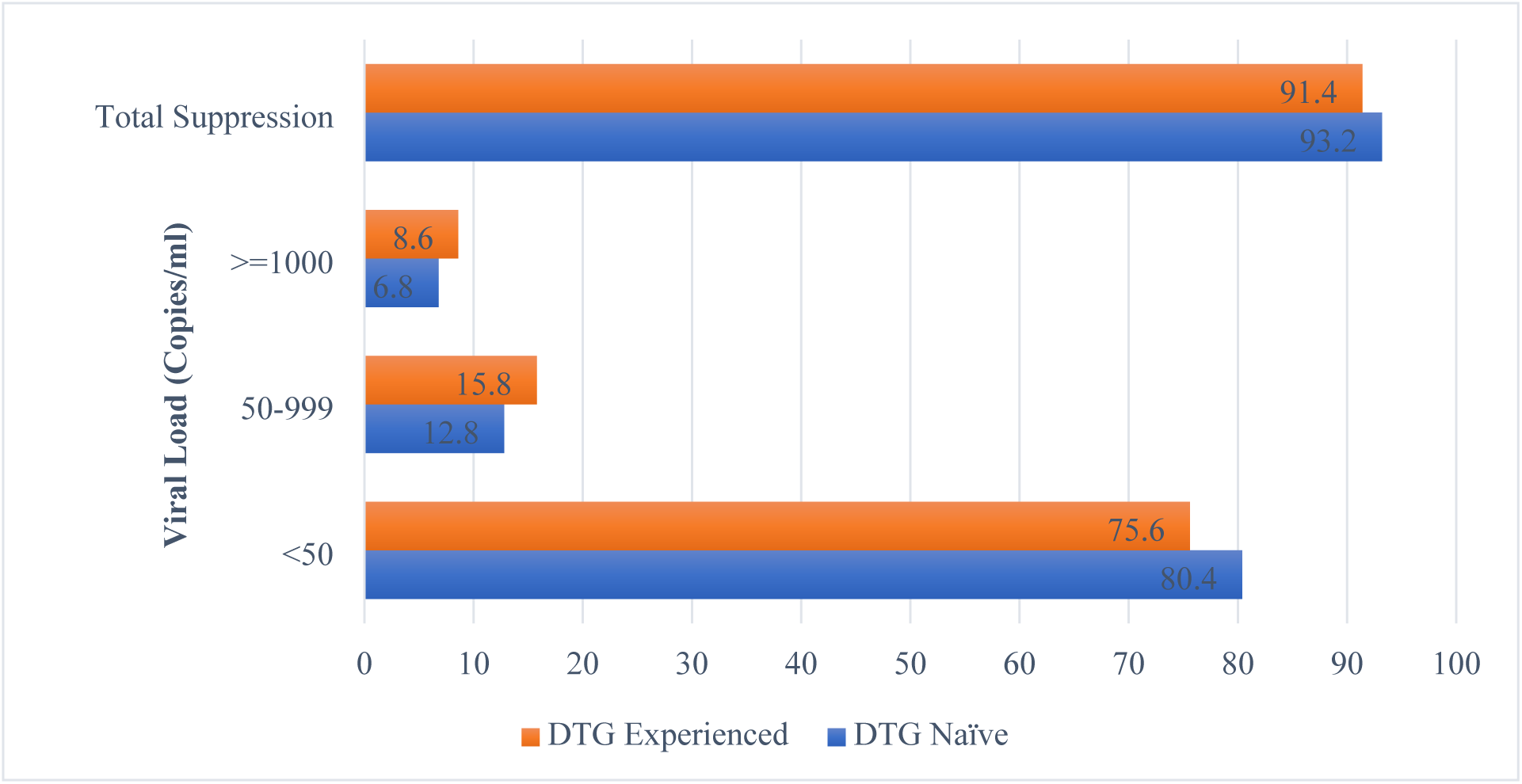
Viral Load among treatment naïve and experienced CALHIV on DTG-based regimen (N=63,453) p-value <0.001.

Among the 63,453 children and adolescents living with HIV on DTG, the prevalence of viral suppression was 91.64% (95%CI: 91.42-91.85; N=58,147). The proportion of viral suppression among females was significantly higher with 92.04% (95%CI: 91.76,92.32) compared to viral suppression among males with 91.10% (95%CI: 90.76,91.43).

Among those with pre- and post-DTG VL, 94.28% (95%CI: 94.04,94.52) of previously suppressed participants remained suppressed, 84.85% (95%CI: 84.22,85.45) of previously unsuppressed became virally suppressed and 93.21% (95%CI: 92.59,93.78) of treatment-naïve participants who started DTG became viral suppressed. Among CALHIV on a DTG-based regimen with 24 months or less duration on ART had a significantly higher proportion of viral suppression with 92.72% (95%CI: 92.19,93.21) compared to individuals with a duration of more than two years (12 months) on ART with 91.44% (95% CI: 91.20,91.67). More results are shown in **Table 2**.

**Table 2:** Proportion of HIV viral suppression by Socio-demographic and Clinical Characteristics of the CALHIV on DTG based ART (N=63,453)

| Variables | Viral Load |  |  | P-value |
| --- | --- | --- | --- | --- |
|  | Unsuppressed | Suppressed (<1000 copies/ml) |  |  |
|  | n (%) | n (%) | 95% CI |  |
|  | 5,306 (8.36%) | 58,147 (91.64%) | 91.42,91.85 |  |
| Sex |  |  |  | <0.001 |
| Male | 2,433 (8.90) | 24,912 (91.10) | 90.76,91.43 |  |
| Female | 2,873 (7.96) | 33,235 (92.04) | 91.76,92.32 |  |
| Age (years) |  |  |  | <0.001 |
| 1-4 | 1,684 (7.71) | 20,168 (92.29) | 91.93,92.64 |  |
| 5-9 | 1,810 (9.05) | 18,199 (90.95) | 90.55,91.34 |  |
| 10-14 | 1,123 (9.59) | 10,584 (90.41) | 89.86,90.93 |  |
| 15-19 | 689 (6.97) | 9,196 (93.03) | 92.51,93.52 |  |
| WHO Staging |  |  |  | <0.001 |
| Stage I | 2,015 (7.74) | 24,011 (92.26) | 91.93,92.58 |  |
| Stage II | 1,585 (8.46) | 17,156 (91.54) | 91.14,91.93 |  |
| Stage III | 1,449 (8.99) | 14,663 (91.01) | 90.56,91.44 |  |
| Stage IV | 254 (9.90) | 2,311 (90.10) | 88.88,91.20 |  |
| CD4 (cells/mm3) |  |  |  | 0.008 |
| <200 | 298 (9.61) | 2,803 (90.39) | 89.30,91.38 |  |
| 200-349 | 175 (9.19) | 1,730 (90.81) | 89.43,92.03 |  |
| >=350 | 2,487 (8.16) | 27,985 (91.84) | 91.53,92.03 |  |
| BMI (Kg/M²) |  |  |  | 0.512 |
| Underweight | 1,610 (8.71) | 16,868 (91.29) | 90.87,91.69 |  |
| Normal | 262 (7.91) | 3,050 (92.09) | 91.12,92.96 |  |
| Overweight | 44 (8.63) | 466 (91.37) | 88.60,93.52 |  |
| Obesity | 119 (8.58) | 1,268 (91.42) | 89.83,92.78 |  |
| Prior DTG status |  |  |  | <0.001 |
| Experienced Suppressed | 1,975 (5.72) | 32,582 (94.28) | 94.04,94.52 |  |
| Experienced Unsuppressed | 1,976 (15.15) | 11,065 (84.85) | 84.22,85.45 |  |
| Naive | 471 (6.79) | 6,462 (93.21) | 92.59,93.78 |  |
| Duration on ART |  |  |  | <0.001 |
| 24 months or less | 724 (7.28) | 9,217 (92.72) | 92.19,93.21 |  |
| >24 months | 4,582 (8.56) | 48,930 (91.44) | 91.20,91.67 |  |
| Current DTG Regimen |  |  |  | 0.382 |
| Other DTG regimen | 109 (7.31) | 1,383 (92.69) | 91.26,93.81 |  |
| TDF+3TC+DTG (TLD) | 389 (6.67) | 5,445 (93.33) | 92.66,93.94 |  |
| Ever Received MMS |  |  |  | <0.001 |
| No | 1,785 (23.88) | 5,691 (76.12) | 75.14,77.08 |  |
| Yes | 3,521 (6.29) | 52,456 (93.71) | 93.51,93.91 |  |
| Child exposed to PMTCT |  |  |  | 0.147 |
| No | 4,896 (8.41) | 53,321 (91.59) | 91.36,91.81 |  |
| Yes | 410 (7.83) | 4,826 (92.17) | 91.41,92.87 |  |
| History of TB treatment |  |  |  | 0.001 |
| No | 4,749 (8.25) | 952,843 (91.75) | 91.53,91.98 |  |
| Yes | 557 (9.50) | 5,304 (90.50) | 89.72,91.22 |  |
| **Malnutrition Status** |  |  |  | <0.001 |
| No Malnutrition | 4,671 (8.40) | 50,930 (91.60) | 91.37,91.83 |  |
| Mild & Moderate | 49 (10.89) | 401 (89.11) | 85.88,91.68 |  |
| Severe | 10 (31.25) | 22 (68.75) | 50.72,82.46 |  |
| **Retention in Care** |  |  |  | <0.001 |
| Retained | 5,183 (8.22) | 57,864 (91.78) | 91.56,91.99 |  |
| Not Retained | 123 (30.30) | 283 (69.70) | 65.05,73.99 |  |
| **Facility type** |  |  |  | 0.021 |
| Dispensary | 1,365 (8.81) | 14,129 (91.19) | 90.73,91.63 |  |
| Health centre | 1,751 (8.47) | 18,914 (91.53) | 91.14,91.90 |  |
| Hospital | 2,172 (8.05) | 24,805 (91.95) | 91.62,92.27 |  |
| **Facility Ownership** |  |  |  | 0.240 |
| Private/FBO | 1,227 (8.14) | 13,839 (91.86) | 91.41,92.28 |  |
| Public | 4,061 (8.45) | 44,009 (91.55) | 91.30,91.79 |  |
| **Zone** |  |  |  | <0.001 |
| Central Zone | 502 (7.71) | 6,008 (92.29) | 91.62,92.91 |  |
| Coastal Zone | 1,352 (10.09) | 12,048 (89.91) | 89.39,90.41 |  |
| Lake Zone | 1,023 (5.96) | 16,129 (94.04) | 93.67,94.38 |  |
| Northern Zone | 657 (8.72) | 6,879 (91.28) | 90.62,91.90 |  |
| Southern Highland | 1,618 (9.57) | 15,289 (90.43) | 89.98,90.86 |  |
| Western Zone | 136 (8.34) | 1,495 (91.66) | 90.22,92.91 |  |

Data are presented as n (row%). Abbreviations: CI, Confidence Interval; n, frequency; %, percentage; DTG, dolutegravir; TDF, tenofovir; 3TC, lamivudine; ABC, abacavir; AZT, azidothymidine (Zidovudine); MMS, Multi-month subscription; FBO, Faith based organizations; ART, Antiretroviral Therapy \*Chi-squared P-value

### Factors associated with viral suppression among CALHIV on DTG-based ART

#### Findings from crude analysis

Females are 1.01 more likely to achieve viral suppression compared to males (cRR: 1.01; 95% CI: 1.01-1.02). Participants who were ART naïve when starting a DTG based regimen are 1.01 more likely to achieve viral suppression compared to participants previously suppressed prior to DTG (cRR: 1.01; 95% CI: 1.00-1.02). Participants who ever received a multi-month subscription are 1.23 more likely to achieve viral suppression compared to participants who never received a multi-month prescription (cRR: 1.23; 95% CI: 1.23-1.25). More results are shown in **Table 3**.

**Table 3:**
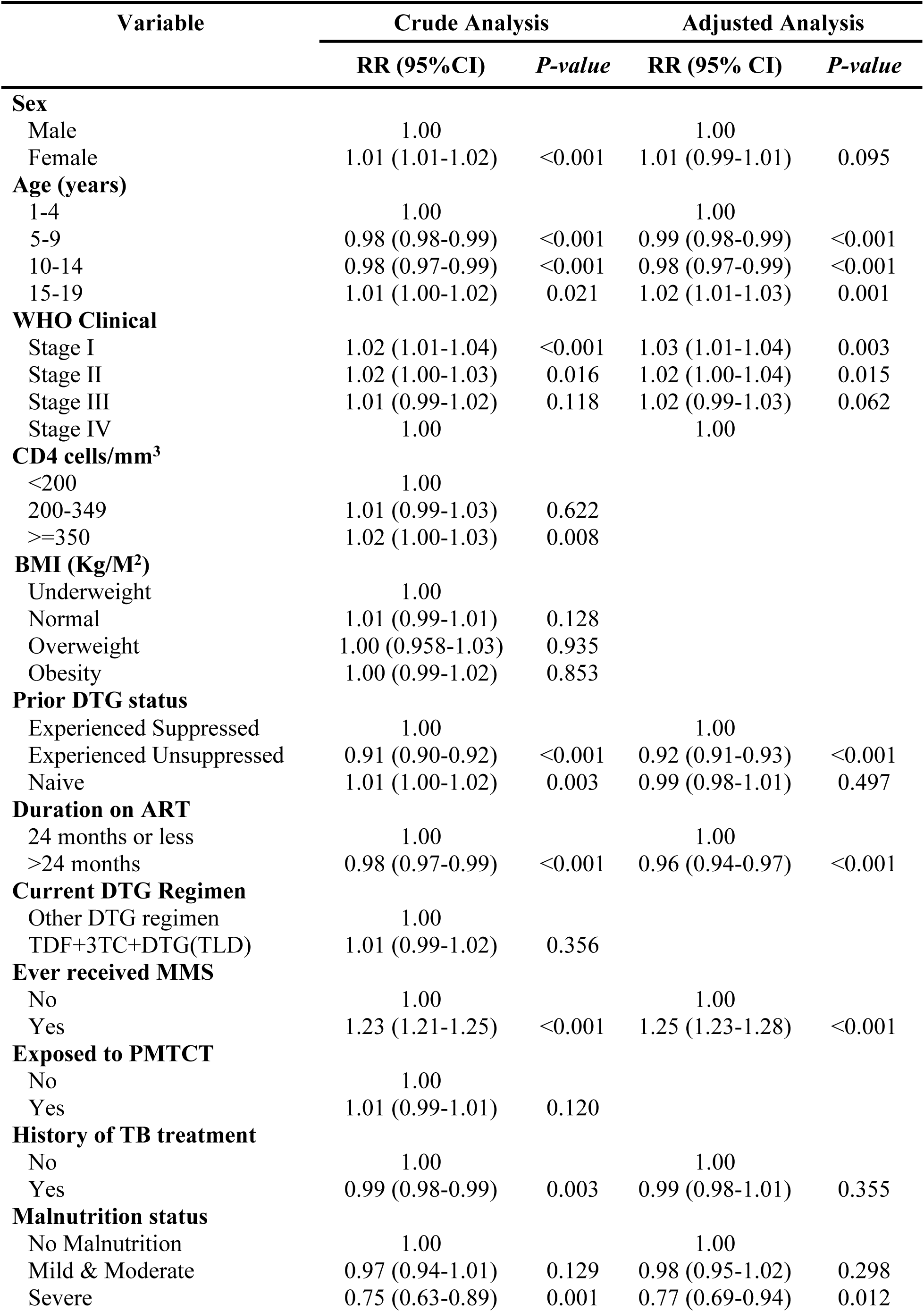
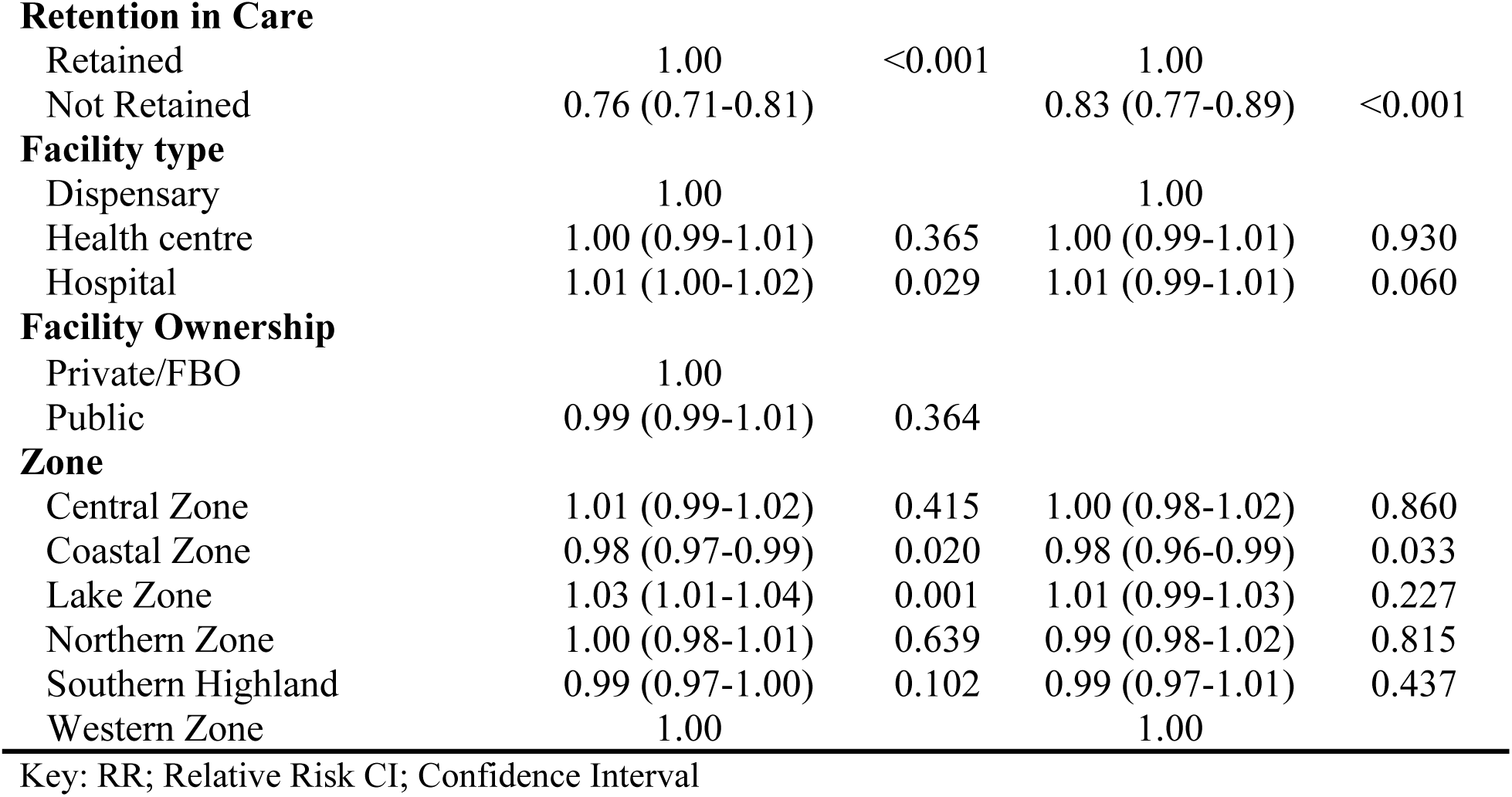
Factors associated with viral suppression among CALHIV on DTG-based ART adjusting for clustering effect (N=63,453)

| Variable | Crude Analysis |  | Adjusted Analysis |  |
| --- | --- | --- | --- | --- |
|  | RR (95%CI) | *P-value* | RR (95% CI) | *P-value* |
| **Sex** |  |  |  |  |
| Male | 1.00 |  | 1.00 |  |
| Female | 1.01 (1.01-1.02) | <0.001 | 1.01 (0.99-1.01) | 0.095 |
| **Age (years)** |  |  |  |  |
| 1-4 | 1.00 |  | 1.00 |  |
| 5-9 | 0.98 (0.98-0.99) | <0.001 | 0.99 (0.98-0.99) | <0.001 |
| 10-14 | 0.98 (0.97-0.99) | <0.001 | 0.98 (0.97-0.99) | <0.001 |
| 15-19 | 1.01 (1.00-1.02) | 0.021 | 1.02 (1.01-1.03) | 0.001 |
| **WHO Clinical** |  |  |  |  |
| Stage I | 1.02 (1.01-1.04) | <0.001 | 1.03 (1.01-1.04) | 0.003 |
| Stage II | 1.02 (1.00-1.03) | 0.016 | 1.02 (1.00-1.04) | 0.015 |
| Stage III | 1.01 (0.99-1.02) | 0.118 | 1.02 (0.99-1.03) | 0.062 |
| Stage IV | 1.00 |  | 1.00 |  |
| **CD4 cells/mm<sup>3</sup>** |  |  |  |  |
| <200 | 1.00 |  |  |  |
| 200-349 | 1.01 (0.99-1.03) | 0.622 |  |  |
| >=350 | 1.02 (1.00-1.03) | 0.008 |  |  |
| **BMI (Kg/M<sup>2</sup>)** |  |  |  |  |
| Underweight | 1.00 |  |  |  |
| Normal | 1.01 (0.99-1.01) | 0.128 |  |  |
| Overweight | 1.00 (0.958-1.03) | 0.935 |  |  |
| Obesity | 1.00 (0.99-1.02) | 0.853 |  |  |
| **Prior DTG status** |  |  |  |  |
| Experienced Suppressed | 1.00 |  | 1.00 |  |
| Experienced Unsuppressed | 0.91 (0.90-0.92) | <0.001 | 0.92 (0.91-0.93) | <0.001 |
| Naive | 1.01 (1.00-1.02) | 0.003 | 0.99 (0.98-1.01) | 0.497 |
| **Duration on ART** |  |  |  |  |
| 24 months or less | 1.00 |  | 1.00 |  |
| >24 months | 0.98 (0.97-0.99) | <0.001 | 0.96 (0.94-0.97) | <0.001 |
| **Current DTG Regimen** |  |  |  |  |
| Other DTG regimen | 1.00 |  |  |  |
| TDF+3TC+DTG(TLD) | 1.01 (0.99-1.02) | 0.356 |  |  |
| **Ever received MMS** |  |  |  |  |
| No | 1.00 |  | 1.00 |  |
| Yes | 1.23 (1.21-1.25) | <0.001 | 1.25 (1.23-1.28) | <0.001 |
| **Exposed to PMTCT** |  |  |  |  |
| No | 1.00 |  |  |  |
| Yes | 1.01 (0.99-1.01) | 0.120 |  |  |
| **History of TB treatment** |  |  |  |  |
| No | 1.00 |  | 1.00 |  |
| Yes | 0.99 (0.98-0.99) | 0.003 | 0.99 (0.98-1.01) | 0.355 |
| **Malnutrition status** |  |  |  |  |
| No Malnutrition | 1.00 |  | 1.00 |  |
| Mild & Moderate | 0.97 (0.94-1.01) | 0.129 | 0.98 (0.95-1.02) | 0.298 |
| Severe | 0.75 (0.63-0.89) | 0.001 | 0.77 (0.69-0.94) | 0.012 |
| **Retention in Care** |  |  |  |  |
| Retained | 1.00 | <0.001 | 1.00 |  |
| Not Retained | 0.76 (0.71-0.81) |  | 0.83 (0.77-0.89) | <0.001 |
| **Facility type** |  |  |  |  |
| Dispensary | 1.00 |  | 1.00 |  |
| Health centre | 1.00 (0.99-1.01) | 0.365 | 1.00 (0.99-1.01) | 0.930 |
| Hospital | 1.01 (1.00-1.02) | 0.029 | 1.01 (0.99-1.01) | 0.060 |

**Facility Ownership**
|  |  |  |
| --- | --- | --- |
| Private/FBO | 1.00 |  |
| Public | 0.99 (0.99-1.01) | 0.364 |
| **Zone** |  |  |  |  |
| Central Zone | 1.01 (0.99-1.02) | 0.415 | 1.00 (0.98-1.02) | 0.860 |
| Coastal Zone | 0.98 (0.97-0.99) | 0.020 | 0.98 (0.96-0.99) | 0.033 |
| Lake Zone | 1.03 (1.01-1.04) | 0.001 | 1.01 (0.99-1.03) | 0.227 |
| Northern Zone | 1.00 (0.98-1.01) | 0.639 | 0.99 (0.98-1.02) | 0.815 |
| Southern Highland | 0.99 (0.97-1.00) | 0.102 | 0.99 (0.97-1.01) | 0.437 |
| Western Zone | 1.00 |  | 1.00 |  |

Key: RR; Relative Risk CI; Confidence Interval

#### Finding from multivariable regression

Participants aged 10-14 years and 5-9 years were 2% less likely to achieve viral suppression compared to participants aged 1-4 years (aRR: 0.98; 95%CI: 0.97-0.99. Also, Individuals in WHO Stage I were 1.03 more likely to achieve viral suppression compared to individuals in WHO stage IV (aRR: 1.03; 95%CI: 1.01-1.04). Participants previously unsuppressed prior to DTG were 8% less likely to achieve viral suppression compared to participants previously suppressed (aRR: 0.92; 95%CI: 0.91-0.93). Participants with a duration on ART of more than 24 months were 4% less likely to achieve viral suppression compared to participants with a duration of 24 months or less (aRR: 0.96; 95%CI: 0.94-0.97). Participants who ever received a multi-month subscription are 1.25 more likely to achieve viral suppression compared to participants who never received a multi-month subscription (aRR: 1.25; 95% CI: 1.23-1.28). Also, those participants not retained in care were 17% less likely to achieve viral suppression compared to participants retained in care (aRR: 0.83; 95% CI: 0.77-0.89). Participants with severe nutritional status were 23% less likely to be HIV virally suppressed compared to participants with no malnutritional nutritional status (aRR:0.77; 95%CI: 0.69-0.94). However, participants living in the coastal zone were 2% less likely to achieve viral suppression compared to the participants living in the western zone (aRR: 0.98; 95% CI: 0.96-0.99). More results are shown in **Table 3**.

## Discussion

This study aimed to determine HIV viral suppression and associated factors among children and adolescents who were on a DTG-based regimen in Tanzania Mainland from 2019 to 2021. The proportion of children and adolescents living with HIV on a DTG-based ART regimen was 41.96% among all CALHIV on ART in Tanzania Mainland. The proportion of viral suppression among CALHIV on DTG based was 91.64%. Furthermore, 66.19% of previously unsuppressed became suppressed and 88.45% of previously suppressed remained suppressed.

The proportion of HIV viral suppression on a DTG-based regimen among children and adolescents living with HIV in our study is approximately similar to 91.5% and 92%, which were reported in Nigeria [10] and Ethiopia [8]. Though, it is higher compared to 84.00% and 80.0% which were reported in the USA [11] and Kenya [12]. A possible explanation is the difference in management guidelines of viral suppression across countries that explains variations based on standard copies/ml per country.

Among previously unsuppressed patients at baseline, 66.1% achieved viral suppression after shifting to DTG-based regimen and among previously suppressed patients at baseline, 88.4% remained suppressed. This figure is approximately similar to studies reported in Italy, Ireland, and Nigeria [9, 10, 11]. Among ART-naïve participants on DTG-based regimen, viral suppression of 91.7% after six months period. This figure is approximately similar to studies in Uganda and France [12, 13]. In addition, ART naïve participants had a higher proportion of viral suppression (93.2%) compared to ART experienced who shifted to a DTG based regimen with 91.4%. The findings are similar to studies in western India [18], Nigeria [14], and Uganda [16]. It showed a good response to the DTG-based regimen which may reduce the risk of developing viral resistance [19].

This study found several factors independently associated with viral suppression including duration on ART of more than 24 months. Similar findings have been reported elsewhere in the USA [20], France [17], and Kenya [12]. It can be explained by the evidence from clinical practice, which suggested that prolonged ART can reduce viral load below the limit of detection for a long-term period [8].

We also found ‘Ever received a multi-month prescription (MMP)’ to be significantly related with viral suppression compared to participants who never received a multi month prescription. Participants who ever received multi-month prescriptions are 1.25 more likely to achieve viral suppression compared to those who never received multi-month prescriptions, which is consistent with the findings of studies conducted in Ethiopia [21] and Kenya [22]. It may be the case that those who do not receive this multi-month prescription for viral suppression have poor adherence due to a drop in drug concentration in body fluids because their viral load increases as a result of their inability to suppress HIV replication, which in turn causes their viral load to increase.

In this study, at baseline, we also found the age of children and adolescents living with HIV to have a significant effect on viral suppression. Children and adolescents living with HIV aged 5-14 years were less likely to achieve viral suppression compared to children and adolescents living with HIV aged 1-4 years. The results are consistent with reports from the USA [23] and Zimbabwe [24]. This can be explained by the fact that adolescents and young adults living with HIV experience multiple barriers to adherence including developmental, physical, emotional, behavioural, and social dynamic changes.

Among those previously on ART, a significant association with being previously unsuppressed prior to DTG was associated with being less likely to achieve viral suppression compared to those previously suppressed prior to DTG. Though, those who were ART naïve when starting DTG were more likely to achieve viral suppression compared to those who were previously suppressed prior to DTG, though not statistically significant. The findings are consistent with the study done in nine countries which involved Africa, Asia, North America, and South America [25].

Furthermore, in this study those not retained in care were less likely to achieve viral suppression compared to CALHIV retained in care, which mirrors the findings of the study done in South Africa [26]. This indicates that patients being retained in care were more likely to achieve viral suppression compared to not retained in regular care since adolescents seem to have lower retention rates compared to other age groups.

This study utilized the national representative data of children and adolescents living with HIV. Hence, enough statistical power increases the precision of the estimates and makes the findings generalizable to treatment naïve and experienced HIV patients. This is the first in Tanzania to evaluate the HIV viral suppression on a dolutegravir-based regimen and associated factors among children and adolescents living with HIV on ART. Therefore, our study has provided an estimate for viral suppression amongst children and adolescents living with HIV on DTG based regimen and associated factors in Tanzania.

This study has a few limitations related to its retrospective nature which utilized secondary data, which might have compromised the data quality and thereby the estimates. This includes missing values in viral load and exposures. The inability to conduct drug resistance testing and antiretroviral drug level testing, which could have helped to determine the contributions of resistance before switching to dolutegravir (DTG) to the risk of treatment failure, especially among virally unsuppressed children and adolescents living with HIV on ART.

## Conclusions

The proportion of viral suppression among children and adolescents living with HIV on dolutegravir (DTG) based regimen in this study was 91.64%. The results encourage widespread use of DTG among eligible CALHIV, especially those in baseline WHO stage I and those who ever received a multi-month prescription, as they were more likely to achieve viral suppression. Viral suppression among CALHIV on DTG was associated with age 5-14, duration on ART more than 24 months, ever received a multi-month prescription, WHO stage I, experienced unsuppressed prior to DTG, not retained in care and living in coastal zone.

Based on the findings from this study, we recommend the following. All children and adolescents living with HIV who are currently receiving other ART (PI or NNRTI) regimens, particularly those who are unsuppressed should switch to a dolutegravir (DTG)-based regimen if eligible. We recommend this because of its strong association in reducing viral load. Dolutegravir based regimen maintains viral suppression but weight gain after its use needs further research to check whether it is beneficial or harmful.

## Authors’ contributions

AAM contributed to the design of the study, performed the statistical analysis, and drafted the manuscript. MSdB, GM, CA and MJM edited the final draft of the manuscript and approved the final manuscript.

## Data Availability

No-some restrictions will apply.

## Acknowledgments

Besides my supervisors, I would like to thank all members of the Department of Epidemiology and Applied Biostatistics and National Aids Control Programme (NACP) for their assistance, encouragement, constructive comments, and recommendation. I do extend my sincere gratitude to the National Aids Control Program (NACP) for the permission to access patient level information in the CTC 2 Databases.

## List of abbreviations

ART: Antiretroviral Therapy
CALHIV: Children and Adolescents Living with HIV
CTC: Care and Treatment Clinic
DTG: Dolutegravir
HIV: Human Immunodeficiency Virus
MMP: Multi-month Prescription
NNRTI: Non-Nucleoside Reverse Transcript Inhibitor
PI: Protease Inhibitor
VS: Viral Suppression
WHO: World Health Organization

